# Hierarchical Clustering with an Ensemble of Principle Component Trees for Interpretable Patient Stratification

**DOI:** 10.1101/2025.02.10.25321980

**Authors:** Bastian Pfeifer, Marcus D. Bloice, Christel Sirocchi, Markus Loecher

## Abstract

Patient stratification plays a crucial role in personalized medicine by identifying distinct subgroups of patients based on their molecular and/or clinical characteristics. However, many unsupervised machine learning-based stratification techniques fail to identify the essential biomarker traits associated with each patient group. In this paper, we present a novel approach for interpretable patient stratification using hierarchical ensemble clustering. Our method leverages feature sampling in conjunction with principal component analysis (PCA) to capture the most significant patterns and contributing biomarkers. We demonstrate the effectiveness of our approach using machine learning benchmark datasets and real-world data from The Cancer Genome Atlas (TCGA), showcasing the improved interpretability of the detected patient clusters.

## 1. Motivation

Clustering for patient stratification or disease subtyping is a valuable strategy that employs computational methods to categorize patients based on shared characteristics, such as clinical features, genetic profiles, or patterns of biomarker expression [7]. By harnessing sophisticated clustering algorithms, researchers can detect distinct subgroups within a disease, leading to a more refined comprehension of its underlying molecular fingerprints [13,10]. This subtyping approach holds significant promise for personalized medicine, as it facilitates tailored treatments and targeted interventions, ultimately improving patient outcomes.

While numerous clustering methods have been developed and applied for patient stratification, less attention has been devoted to making such unsupervised models interpretable, for instance, by identifying the features most relevant for assigning patients to each cluster. Algorithms to verify feature importance contribute to interpretable machine learning by providing insight into how models make decisions [4]. By identifying which features have the greatest impact on model predictions, these algorithms help clarify the relationships between input data and outcomes. This transparency allows practitioners to validate the model’s behavior and detect biases. Moreover, feature importance analysis enhances trust and accountability in machine learning, making it easier to understand and justify the model’s predictions, especially in critical applications like healthcare [11]. However, feature importance has predominantly been explored in supervised machine learning, while it has received less attention in unsupervised settings such as clustering. This proposed approach contributes to addressing this gap.

In disease subtyping, such investigations could reveal that certain risk groups exhibit a higher relevance of particular genes or biomarkers, underscoring their importance in understanding the risk associated with those subgroups. Identifying and prioritizing these distinctive genetic factors within clusters is crucial for targeted interventions and personalized risk assessments. The methodologies presented in this study aim to rectify this gap in previous research by elucidating the features driving patient stratification for enhanced clustering interpretability.

The remainder of the paper is organized as follows: section 2 introduces our new approach to interpretable disease subtyping. Section 2.1 presents the algorithmic details of our ensemble clustering technique, section 2.2 describes the capacity of our approach to handle multi-omics data. A possible extension for longitudinal data is discussed in section 2.3, while section 2.4 outlines our novel strategy to compute cluster-specific feature importance values. In section 3 we describe our evaluation strategy and the results are discussed in section 4. We conclude with an outlook for future work in section 5.

## 2. New Approach

The proposed methodology is called TAPIO (**T**ree **A**ffinities with **P**r**i**ncipal C**o**mponents) and consists of two novel developments. First, instead of performing clustering on the entire dataset, including the full set of features, we suggest that a certain amount of features are sampled without replacement and to compute the first principal component based on this data subset. Hierarchical clustering is then executed on this embedded single feature to construct a dendrogram, which ultimately is split at multiple levels to derive multiple cluster partitions. These partitions are utilized to generate an affinity matrix. This process is iterated multiple times, resulting in an ensemble of affinity matrices, which are aggregated into a single matrix on which clustering is performed. Second, based on the above described scheme, we evaluate cluster-specific feature relevance by computing the contribution of each feature to the principal components. For each ensemble member, the feature contribution to the principal component is scaled by the pair-wise affinity of the elements of the considered cluster. Subsequently, these contributions are aggregated across ensemble members.

### 2.1 Hierarchical Ensemble Learning

Let’s denote the input tabular data as *X*, where *X* is an *n × p* matrix, with *n* samples (patients) and *p* features. Each row of *X* represents a patient, and each column represents a feature. Our developed algorithm consists of five steps:

1. Feature Sampling: Randomly select a subset of *k* features from the dataset *X* to construct a new *n× k* matrix termed *X*^*′*^.
2. Principal Component Analysis (PCA): Reduce the dimensionality of the matrix *X*^*′*^ via PCA. This entails performing eigendecomposition of the covariance matrix 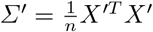 to obtain its eigenvectors *v*_*i*_ and corresponding eigenvalues *λ*_*i*_ (with *i* ranging from 1 to *k*), selecting the *q* eigenvectors corresponding to the *q* largest eigenvalues to form the matrix *V*_*q*_, and projecting the matrix *X*^*′*^ onto the subspace spanned by the first *q* principal components: *Z* = *X*^*′*^*V*_*q*_. Here, *Z* is the *n× q* matrix of the first *q* principal components of *X*^*′*^. In this context, *q* = 1 as only the principal component is considered.
3. Hierarchical Clustering: Apply hierarchical clustering on the reduced data *Z*. Let *D* denote the resulting dendrogram obtained from hierarchical clustering.
4. Dendrogram Cutting: Cut the dendrogram *D* at the first *d* levels, resulting in a set of *d* partitions *C*^*e*^, with *e* ranging from 1 to *d*, where each partition *C*^*e*^ = {*C*_1_, *C*_2_, .., *C*_*p*_} assigns each of the *n* data samples to a cluster *C*. Then, *C*^*e*^(*i*) = *C*_*f*_ indicates that, at level *e*, patient *i* is assigned to the cluster C_f_ ∈ *C* ^*e*^.
5. Affinity Matrix Construction: Construct the *n× n* affinity matrix *A* based on the resulting partitions, such that each entry *A*_*ij*_ represents the strength of the affinity between patient *i* and patient *j* across the partitions. This is computed as the proportion of samples that fall in the same cluster:

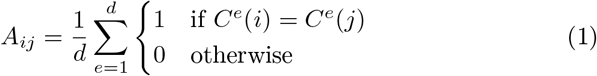

The above described procedure is executed *M* times for the construction of an ensemble consisting of *M* affinity matrices ***A*** = {*A*^(1)^, *A*^(2)^, …, *A*^(*M*)^}. We aggregate these affinity matrices into a single matrix *A* using element-wise summation:

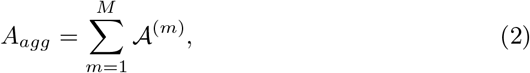

The aggregated affinity matrix *A*_*agg*_ is clustered to obtain the final partitioning of the data.

### 2.2. Integrating Multi-Omics Data

The availability of multiple omics datasets for the same group of patients offers the potential to improve patient stratification, enabling the identification of more precise molecular subtypes. Here, we utilize Multiple Factor Analysis (MFA) when multi-omics data is available. MFA is a direct and straightforward extension of PCA to accommodate multiple data types. The key algorithmic extension lies in a pre-processing step of matrix *X*^*′*^. Let’s assume *X*^*′*^ consists of a list of termed input matrices 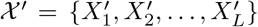, where *L* denotes the total number of available omic data tables. Balance among these data types is achieved by normalizing each of the data types in χ^*′*^, before stacking them and passing them on to PCA. The normalization is accomplished by dividing the principal component decomposition of each termed matrix by its first eigenvalue:

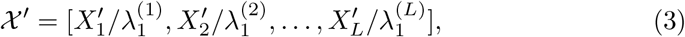

where 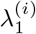 is the first eigenvalue of the principal component decomposition of 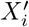, and *i* ∈ *{*1, 2, …, *L*}. Following this normalization and column-wise concatenation step, we apply PCA to *χ*^*′*^ and proceed as described in section 2.1.

### 2.3 Longitudinal Data

In longitudinal studies, variables are measured repeatedly over time, resulting in observations that are both clustered and correlated. This type of data structure, known as repeated measures data sets, is prevalent not only in medical research [15] but also in other fields, such as marketing. For instance, marketing research often involves weekly household purchase data and responses to panel survey questionnaires [1]. Clustering longitudinal data presents unique challenges due to the necessity of grouping based on the similarity of individual trajectories while accommodating sparse and irregular times of observation [19]. These complexities underscore the importance of developing effective methods for clustering longitudinal data, which can provide valuable insights across various domains.

In [3], the *KmL3D* software package was introduced, employing a specialized version of k-means to cluster joint trajectories. The distance metric between individuals is defined as the Minkowski distance between their respective trajectory matrices. [19] introduces a hierarchical agglomerative clustering approach that uses a dissimilarity metric to quantify the merging cost between two groups of curves, represented by B-splines for repeatedly measured data.

None of the outlined methods can be applied to our setting in a straightforward manner. Instead, we propose to add one more sampling step to the algorithm described in section 2.1 to handle longitudinal data:

At each iteration we sample not only *k* features but also one row (time of measure) from each patient. For sufficiently large numbers of *M* iterations we would actually “cover” all the multiple time trajectories from each user. In this way, the clustering algorithm would take the entire variability of each user into account and not just the first moment. It is evident that averaging replicated data for each subject, or even the first principal component, results in a loss of information as intra-subject variation is effectively smoothed out. The most similar approach to our method that we found in the literature is the “subject-level bootstrapping strategy” [6,5], where bootstrap resampling is conducted at the subject level, and all observations from the chosen subjects are treated as in-bag samples.

Implementing this proposed strategy would be beyond the scope of this paper.

### 2.4 Feature Importance

Feature importance is derived from the feature contribution to the principal components used in each ensemble member. In PCA, the loading matrix **V** contains the loading vectors as its columns. In the case of *p* original features and *q* principal components, then **V** is a *p × q* matrix where each column represents the loading vector for a principal component. Within the constructed ensemble, **V**^(*m*)^ is the loading matrix obtained for the *m*^*th*^ member of the ensemble. In this context, *q* = 1 as only the principal component is considered, so **V** is a *p ×* 1 vector. Then, for a specific cluster *cl* the feature importance *ℐ*_*cl*_ is the *p×* 1 vector, with the *i*^*th*^ element of the vector corresponding to the cluster-specific importance of the *i*^*th*^ feature of the dataset, computed as follows:

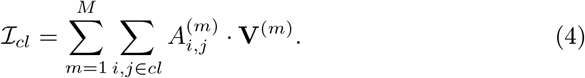

The values in *ℐ*_*cl*_ are normalized by the maximum value across all cluster-specific importance values **max**_*i*,*j*_ 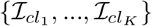, where *K* refers to the number of clusters.

## 3 Evaluation Strategy

Our evaluation approach was two-fold. First, we have validated the proposed methods based on six machine learning datasets (see Table 1), in terms of accuracy and cluster interpretability. Since we have the ground-truth in these cases we could asses the performance of TAPIO utilizing the Adjusted Rand Index (ARI). We performed the clustering with Ward’s linkage [18,9] 100 times for each set-up to study the robustness while varying the number of trees within the ensemble. The number of features randomly sampled for building a dendrogram was set to 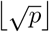. The number of levels to cut the dendrograms was set to 30. We compared the results to a baseline, where Ward’s linkage clustering is applied to the full dataset. Furthermore, we have computed feature importance values for each variable within each cluster according to equation 4.

**Table 1.**
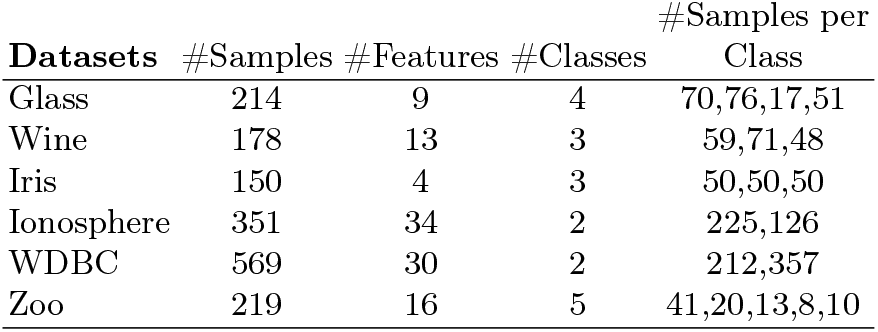
Benchmark data sets.

| **Datasets** | **#Samples** | **#Features** | **#Classes** | **#Samples per Class** |
| --- | --- | --- | --- | --- |
| Glass | 214 | 9 | 4 | 70,76,17,51 |
| Wine | 178 | 13 | 3 | 59,71,48 |
| Iris | 150 | 4 | 3 | 50,50,50 |
| Ionosphere | 351 | 34 | 2 | 225,126 |
| WDBC | 569 | 30 | 2 | 212,357 |
| Zoo | 219 | 16 | 5 | 41,20,13,8,10 |

Second, we retrieved real-world gene expression data from The Cancer Genome Atlas (TCGA) and compared TAPIO to three state-of-the-art disease subtyping methods, SNF [17], NEMO [14], and HCFUSED [12]. We conducted a comparison with the competing approaches based on gene expression data. The data table consisted of 208 patients and gene expression values from 20,531 genes. We randomly subsampled 100 patients from the data pool and assessed the performance using the c-index [16] and the log-rank test statistic. This evaluation procedure was repeated 30 times. Additionally, we executed the aforementioned evaluation procedure using multi-omics data. Along with gene expression levels, we incorporated DNA methylation data, consisting of 5,000 markers, and microRNA data, which included 1,046 biological features.

Finally, we performed patient stratification on the full dataset and analyzed feature importance for the detected risk clusters.

## 4 Results and Discussion

The evaluation based on the machine learning benchmark datasets underscores the potential of feature sampling in conjunction with Principal Component Analysis (see Figure 1). In five out of six cases TAPIO outperforms the baseline approach. We could show that increasing the number of trees within the ensemble has an overall stabilizing effect on the clustering. For each detected cluster we display the cluster-specific feature importance values, allowing researchers to leverage the key features for further in-depth analysis. As can be seen from Figure 1, the feature importance values can differ substantially among the clusters.

**Fig. 1.**
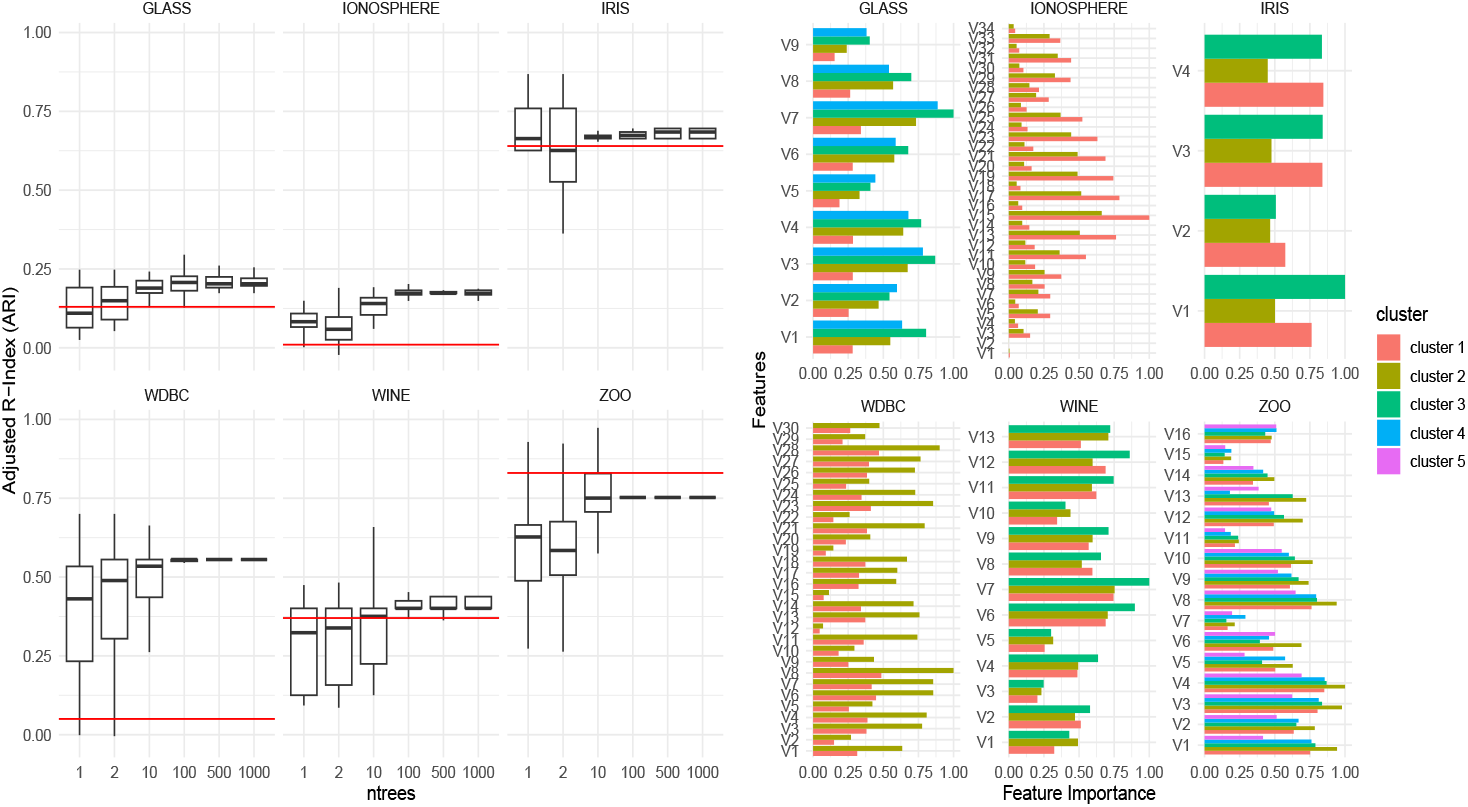
Varying the number of trees. *Left panel*: The ARI performance of our method is displayed for six machine learning benchmark datasets while subsequently increasing the number of trees. The horizontal red lines refer to the performance of the baseline approach. *Right panel*: The cluster-specific feature importance values for each of the six analyzed machine learning benchmark datasets are displayed.

The application of our approach on real-world kidney cancer data revealed competitive results when compared to the state-of-the-art in detecting disease subtypes. Evaluation limited to gene expression revealed log-rank p-values below the significance level for all studied methods (see Figure 2). When integrating multi-omics data, however, TAPIO outperforms the competing methods (see Figure 3), and almost all sub-sampling iterations result in significant log-rank p-values. An increase in performance can also be observed for HCFUSED. Interestingly, NEMO performs worse when multi-omics data is incorporated, compared to using gene expression only (Figure 2).

**Fig. 2.**
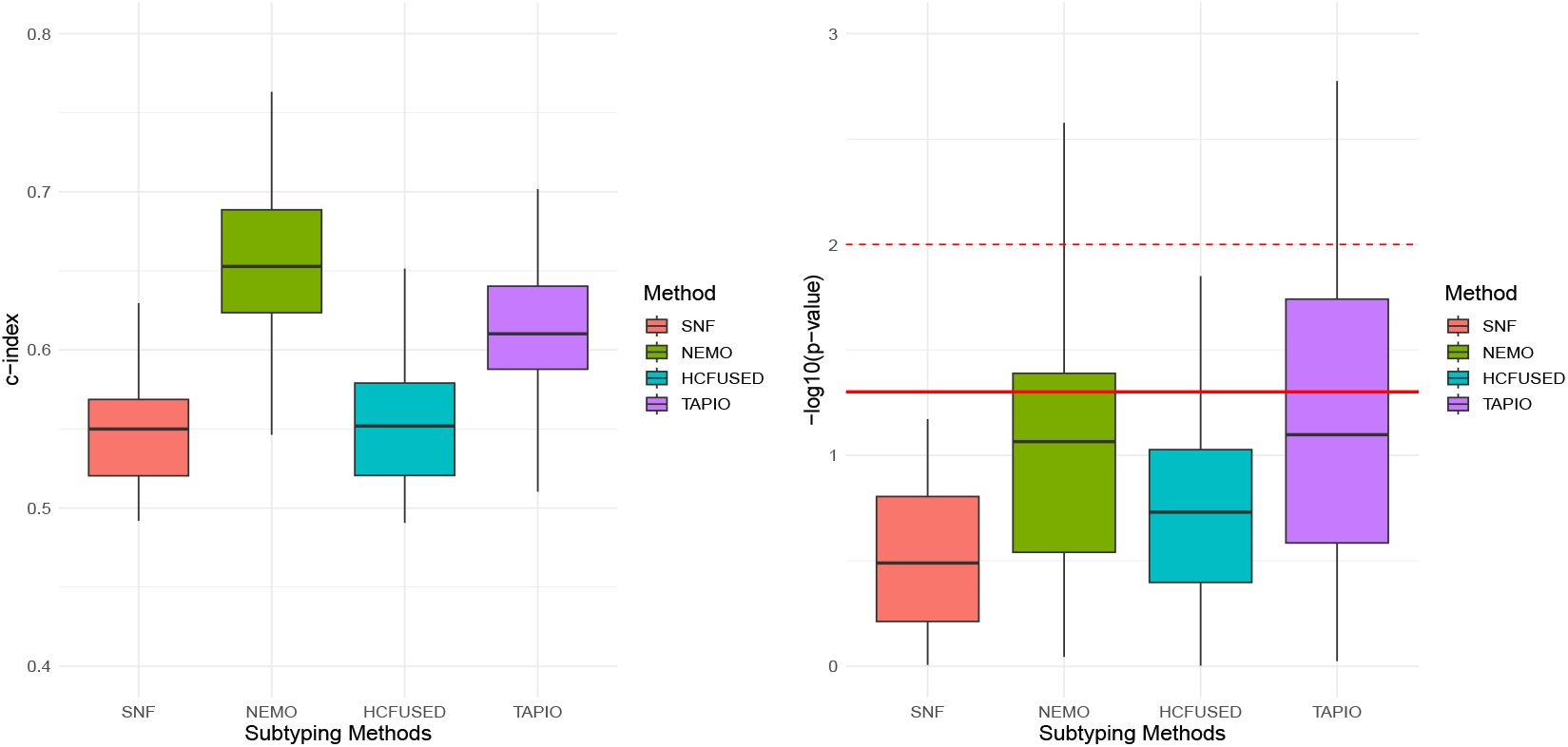
Application on TCGA kidney cancer gene expression data. Our clustering approach TAPIO in comparison with three state-of-the-art disease subtyping methods, NEMO, HCFUSED, and SNF. *Left panel*: Results based on the c-index are displayed. *Right panel*: The log-rank p-values on a logarithmic scale. The red line refers to the 0.05 significance level; the dashed red line refers to the 0.01 significance level. Values above these lines are significant.

**Fig. 3.**
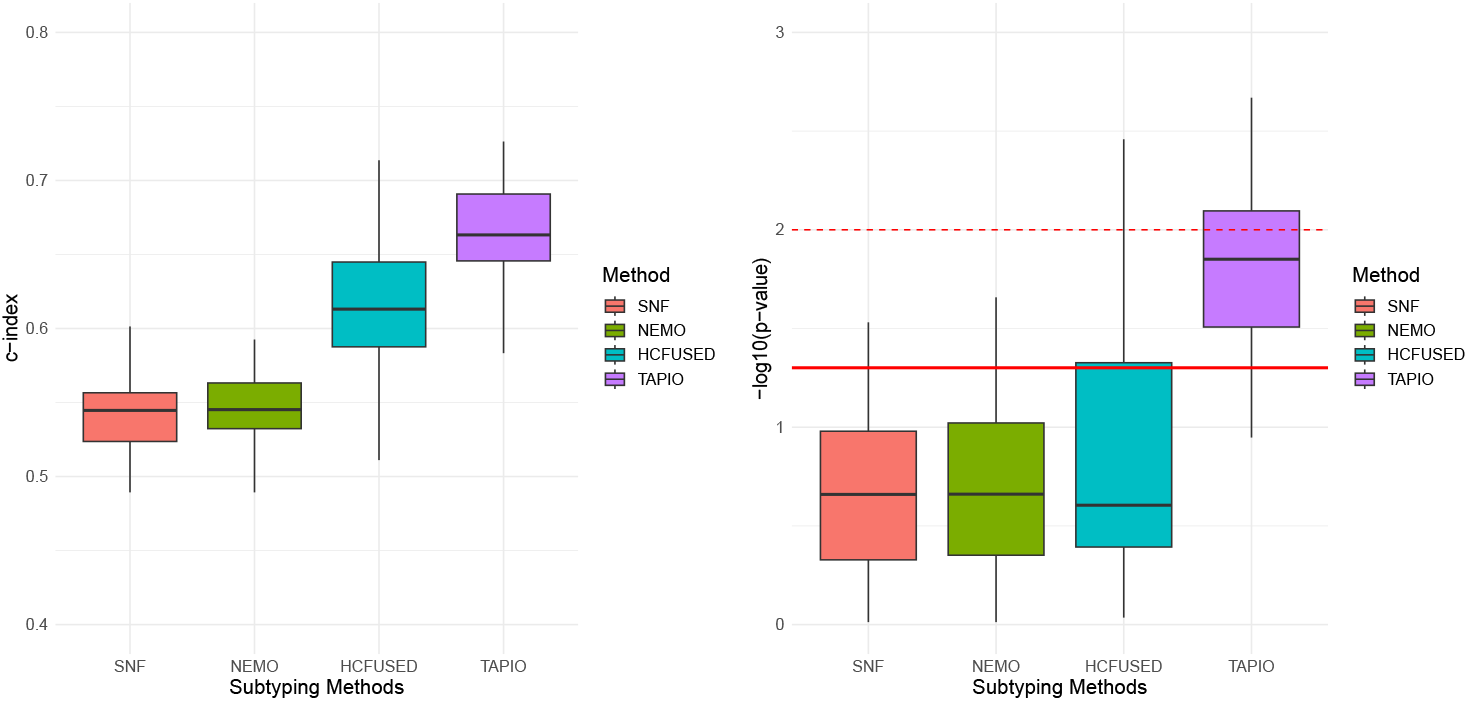
Application on TCGA kidney cancer multi-omics data. Our clustering approach TAPIO in comparison with three state-of-the-art disease subtyping methods, NEMO, HCFUSED, and SNF. *Left panel*: Results based on the c-index are displayed. *Right panel*: The log-rank p-values on a logarithmic scale. The red line refers to the 0.05 significance level; the dashed red line refers to the 0.01 significance level. Values above these lines are significant.

We also assessed TAPIO on liver cancer, and the results presented in Figure 4 demonstrate that TAPIO performs competitively with HCFUSED, while outperforming NEMO and SNF. In general, both TAPIO and HCFUSED appear to perform substantially better on kidney cancer compared to liver cancer. However, TAPIO provides techniques for calculating cluster-specific feature importance, whereas HCFUSED focuses solely on reporting the contribution of a specific molecular modality to the clustering solution. Future work could combine these two approaches to further improve cluster interpretability.

**Fig. 4.**
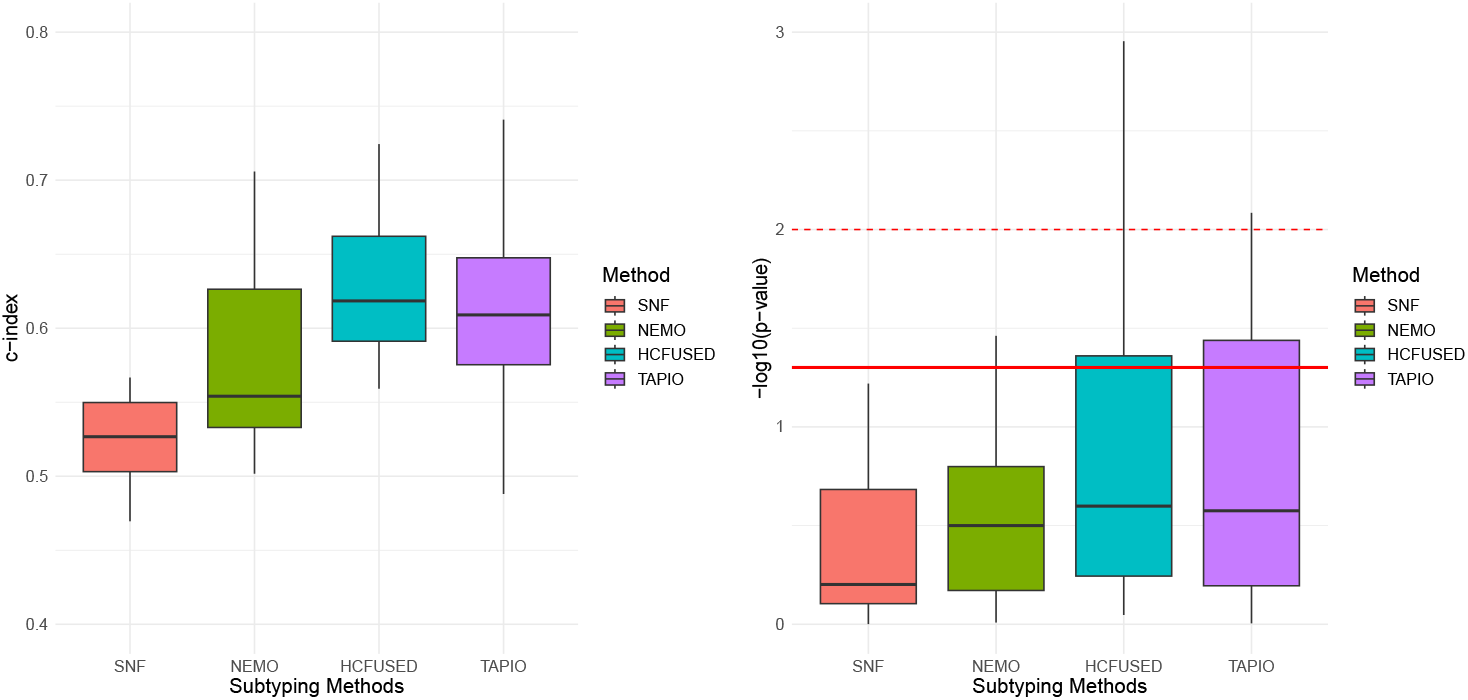
Application on TCGA liver cancer multi-omics data. Our clustering approach TAPIO in comparison with three state-of-the-art disease subtyping methods, NEMO, HCFUSED, and SNF. *Left panel*: Results based on the c-index are displayed. *Right panel*: The log-rank p-values on a logarithmic scale. The red line refers to the 0.05 significance level; the dashed red line refers to the 0.01 significance level. Values above these lines are significant.

Finally, we performed patient stratification on the full kidney cancer dataset using TAPIO. We detected four clusters of size [cl1 = 64, cl2 = 55, cl3 = 73, cl4 = 16] patients with a significant log-rank p-value of *p* = 0.0053. The corresponding survival curves are displayed in Figure 5. Clusters 3 and 4 are high risk clusters with substantially lower survival rates than patient clusters 1 and 2.

**Fig. 5.**
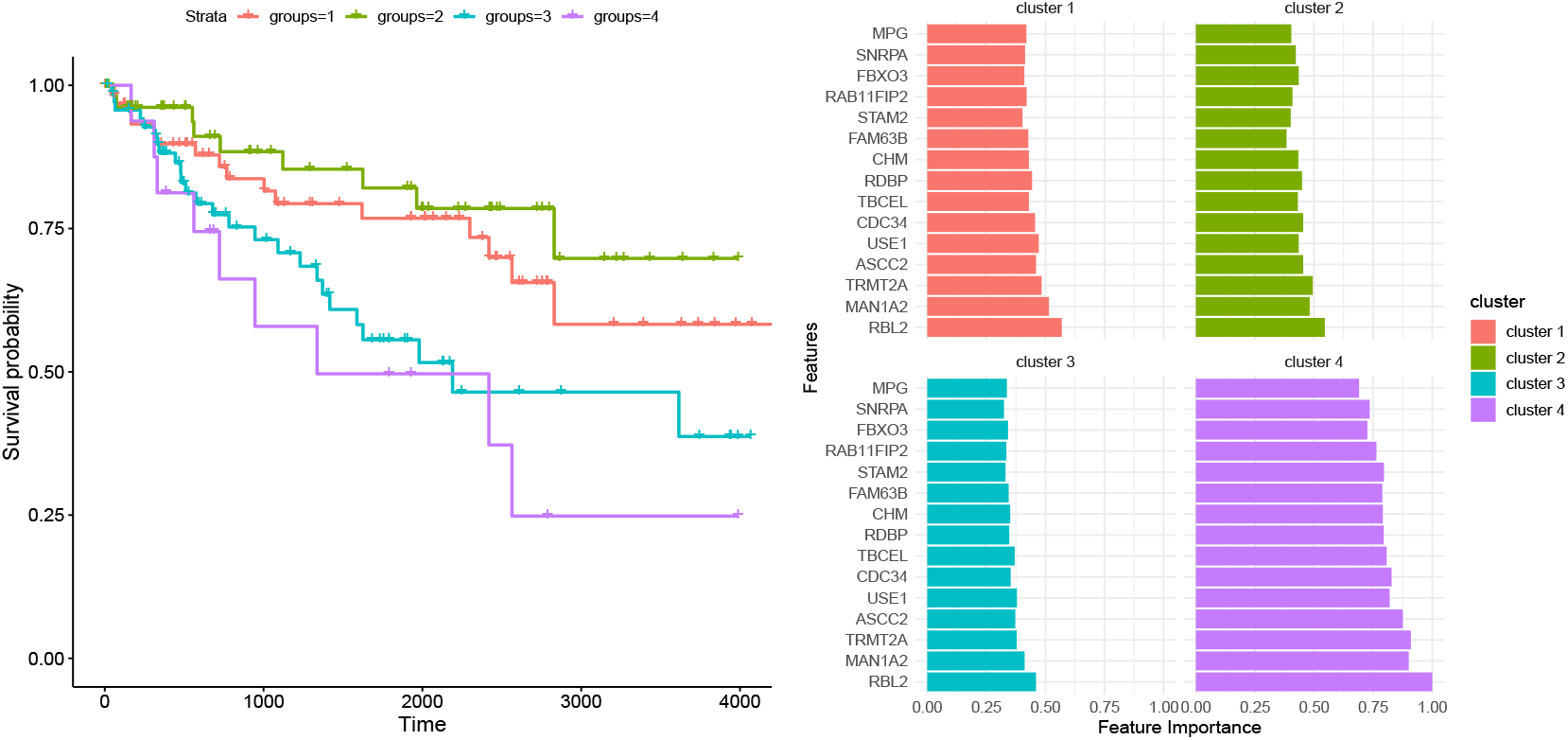
Application on TCGA kidney cancer gene expression data. *Left panel*: Survival curves of the four detected patient groups based on the full kidney cancer dataset. The log-rank p-value is *p* = 0.0053. *Right panel*: Feature importance values of each detected patient group from a selected set of the 15 most relevant genes

Furthermore, we calculated the 15 relevant genes based on the mean across all cluster-specific feature importance values. The gene “RBL2” emerged as the most relevant, consistently ranking as the top priority across all patient clusters. “RBL2” is structurally similar to the gene “RB1”, which is a known tumor suppressor gene. However, it is not yet fully investigated whether it functions in a similar way, thereby providing an opportunity for further investigation [2]. The results shown in Figure 5 contain some interesting results. For high-risk cluster 4, the 15 most relevant genes exhibit significantly elevated importance values relative to the other clusters, suggesting a distinct and crucial function within this particular cluster. In addition, we noted slight variations in the distribution of importance levels among the selected genes within clusters 1 to 3. However, to derive final and medically relevant conclusions further in-depth analyses of the detected genes is required.

## 5. Summary and Future Work

We have presented a novel clustering algorithm for enhancing the interpretability in patient stratification. We could show superior performance of our method over a baseline approach based on six machine learning benchmark datasets. The application on real-world cancer data revealed competitive results in patient stratification when compared to the state-of-the-art. Overall, the interpretability of the obtained clusters could be substantially improved.

In future work, the use of UMAP (Uniform Manifold Approximation and Projection) [8] could be explored as an alternative to PCA for dimensionality reduction. While PCA effectively captures linear relationships and global variance, UMAP has the advantage of preserving both local and global structures in nonlinear data. Investigating UMAP’s potential may enhance the understanding of the detected disease subtypes and improve downstream tasks that rely on the reduced feature space.

Moreover, we plan to incorporate the fusion algorithm implemented in HCFUSED. As a consequence, TAPIO will not only provide insights into the importance of the analyzed features, but also will report on the contribution of each omic type to the final clustering. Furthermore, we intend to analyze the dynamic nature of these clusters over time, considering longitudinal data to capture temporal changes in patient subtypes and disease progression. This longitudinal perspective will offer valuable insights into the evolution of disease subtypes and the effectiveness of interventions tailored to specific cluster profiles, thus advancing our understanding of personalized medicine’s potential impact on long-term patient outcomes.

## Data Availability

All data produced are available online at http://linkedomics.zhang-lab.org/#/ and https://github.com/pievos101/TAPIO.

## Acknowledgments

This work has been funded by the European Union’s Horizon Europe research and innovation programme as part of the PoCCardio project, grant no. 101095432.

## Availability of data and software code

Our software code and the analyzed datasets are available at https://github.com/pievos101/TAPIO.

